# Evaluating the Efficacy of Stay-At-Home Orders: Does Timing Matter?

**DOI:** 10.1101/2020.05.30.20117853

**Authors:** Alexandra Medline, Lamar Hayes, Katia Valdez, Ami Hayashi, Farnoosh Vahedi, Will Capell, Jake Sonnenberg, Zoe Glick, Jeffrey D. Klausner

## Abstract

**BACKGROUND:** The many economic, psychological, and social consequences of pandemics and social distancing measures create an urgent need to determine the efficacy of non-pharmaceutical interventions (NPIs), and especially those considered most stringent, such as stay-at-home and self-isolation mandates. This study focuses specifically on the efficacy of stay-at-home orders, both nationally and internationally, in the control of COVID-19.

**METHODS:** We conducted an observational analysis from April to May 2020 and included countries and US states with known stay-at-home orders. Our primary exposure was the time between the date of the first reported case of COVID-19 to an implemented stay-at-home mandate for each region. Our primary outcomes were the time from the first reported case to the highest number of daily cases and daily deaths. We conducted simple linear regression analyses, controlling for the case rate of the outbreak.

**RESULTS:** For US states and countries, a larger number of days between the first reported case and stay-at-home mandates was associated with a longer time to reach the peak daily case and death counts. The largest effect was among regions classified as the latest 10% to implement a mandate, which in the US, predicted an extra 35.3 days to the peak number of cases (95 % CI: 18.2, 52.5), and 38.3 days to the peak number of deaths (95 % CI: 23.6, 53.0).

**CONCLUSIONS:** Our study supports the potential beneficial effect of earlier stay-at-home mandates, by shortening the time to peak case and death counts for US states and countries. Regions in which mandates were implemented late experienced a prolonged duration to reaching both peak daily case and death counts.

## Background

The coronavirus disease 2019 (COVID-19) is an acute respiratory disease officially declared as a “public health emergency of international concern” by the World Health Organization (WHO) on January 30, 2020.^1^ Since the first case announced on December 8, 2019, in Wuhan, China, COVID-19 has spread internationally with the eventual announcement of a global pandemic by the WHO on March 11, 2020. Healthcare systems and governments worldwide have been under pressure since this designation to implement strategies and containment measures against COVID-19, an unprecedented virus with challenges in all that is left to learn.^2^

Extrapolation from epidemiological models of COVID-19 has suggested that intensive physical distancing could “flatten the curve” and prevent the overloading of our health systems.^3^ Social distancing measures, aimed at reducing contact between people, include school closings, stay-at-home mandates, and government support for telecommuting,^4,5^ and have become commonly adopted practices on a world-wide scale.^6^ These measures aim to reduce the frequency of physical contact between persons, thereby reducing the risk of the spread of COVID-19, which is known to be transmitted through respiratory droplets.^7^ Various degrees of these social distancing measures were employed in the mitigation of previous respiratory viral pandemics such as the 1918-19 influenza pandemic and the 2003 SARS outbreak, when clear pharmaceutical treatments or vaccines were unavailable. Although retrospective reviews of these overarching measures suggest overall unestablished efficacy in quelling the spread of disease,^5^ the challenges and impracticality of imposing these measures have long been acknowledged.^4,8^ Given the devastating economic, psychological, and social consequences associated with pandemics in general^9^ and with COVID-19 specifically,^10,11^ there is a need to clearly distinguish between the efficacy of different social distancing measures, and in particular, the efficacy of those considered most stringent such as stay-at-home and self-isolation mandates.

Pan et al. sought to evaluate the effectiveness of non-pharmaceutical interventions (NPIs) and found that a series of various public health interventions were temporally associated with the improved control of the COVID-19 outbreak in Wuhan, China.^12^ Furthermore, their study concluded that the implementation of NPIs was associated with a reduction of the effective reproductive number (R*t*), defined as the average number of secondary cases per primary case at calendar time *t ^13^*, to below 1.0 on February 6, 2020 and to below 0.3 on March 1, 2020.^12^ Since then, many studies aimed at determining the efficacy of social distancing, mostly within the US, have demonstrated the protective effects of these measures on controlling the spread of COVID-19.^14,15^

The objective of this current study is to add to the growing evidence base on this topic by evaluating the relationship between country- and US state-level stay-at-home orders and the spread of COVID-19. We quantify the time interval between a country or state’s first reported case of COVID-19 and its implementation of a stay-at home-order to assess any relationship with the time between the first reported case and peak burden of COVID-19, measured by both peak daily cases and deaths.

## Methods

### Source of Data

We conducted an observational study from April 2020 to May 2020. First, we collected available retrospective data online from select English-language Ministry of Health and local news websites regarding the date of implementation for stay-at-home orders in countries and US states included in the study. Google was used as our primary search engine. Specific terms used in our online searches included ‘date of stay-at-home orders 2020,’ ‘non-pharmaceutical interventions COVID-19,’ and ‘stay-at-home mandates.’ We conducted a search for each respective country and US state analyzed in the study.

For case and death counts for US states, we used official public health department websites for each respective state. For country-level data, we used WHO daily COVID-19 situation reports as well as data presented on *worldometer.com*.

### Case Definitions and Analysis

Stay-at-home orders were defined as regionwide restrictions of non-essential internal movement (commonly referred to as “lockdowns”).^16^ To assess the efficacy of stay-at-home orders, we measured the number of days between the implementation of a regional stay-at-home order and objective measures of the peak COVID-19 burden for each country and US state. We chose two main outcome variables to reflect this peak, which included: 1. Highest daily case count, 2. Highest daily death count. The highest daily case count was defined as the largest number of laboratory-confirmed cases and the highest daily death count as the largest number of new deaths attributed to COVID-19 per day.

Our primary exposure was the number of days between the first reported case of COVID-19 in a studied area and the date of nation- or state-wide restriction of internal movement. This variable was measured as both a continuous and categorical variable. Each location, based on the number of days between its first case and its stay-at-home mandate, was categorized into one of three equal terciles: early, middle, or late, analyzed with the creation of dummy variables. In addition, based on the frequency distribution for both countries and US states, the earliest and latest 10% to implement mandates were also formed into their own categories. Our primary outcome variables were the number of days from the first reported case of COVID-19 to the peak of daily cases and to the peak of daily deaths, in each respective country and US state included in our analysis.

We conducted simple linear regression analyses, controlling for the regional case rate of the outbreak which was defined as the number of new cases per 100,000 persons, on the day that the mandate was implemented. The analysis was conducted for both included countries and US states. We used SPSS version 26 for our analysis with a significance level of .05.

## Results

### US State-Level Descriptive Analysis

Forty-two states with stay-at-home orders and the state of Kentucky, which implemented a “healthy-at-home” mandate, were included in our analysis. There were eight states without stay-at-home orders that were excluded from our analysis: Arkansas, Iowa, Nebraska, North Dakota, South Dakota, Oklahoma, Utah, and Wyoming. Of the 43 states included, the number of days between the first reported case and the stay-at-home mandate ranged from 7 to 62 days (Fig. 1), with a mean of 24.0 days and a standard deviation of 11.5 days (Fig. 2).

**Figure 1:**
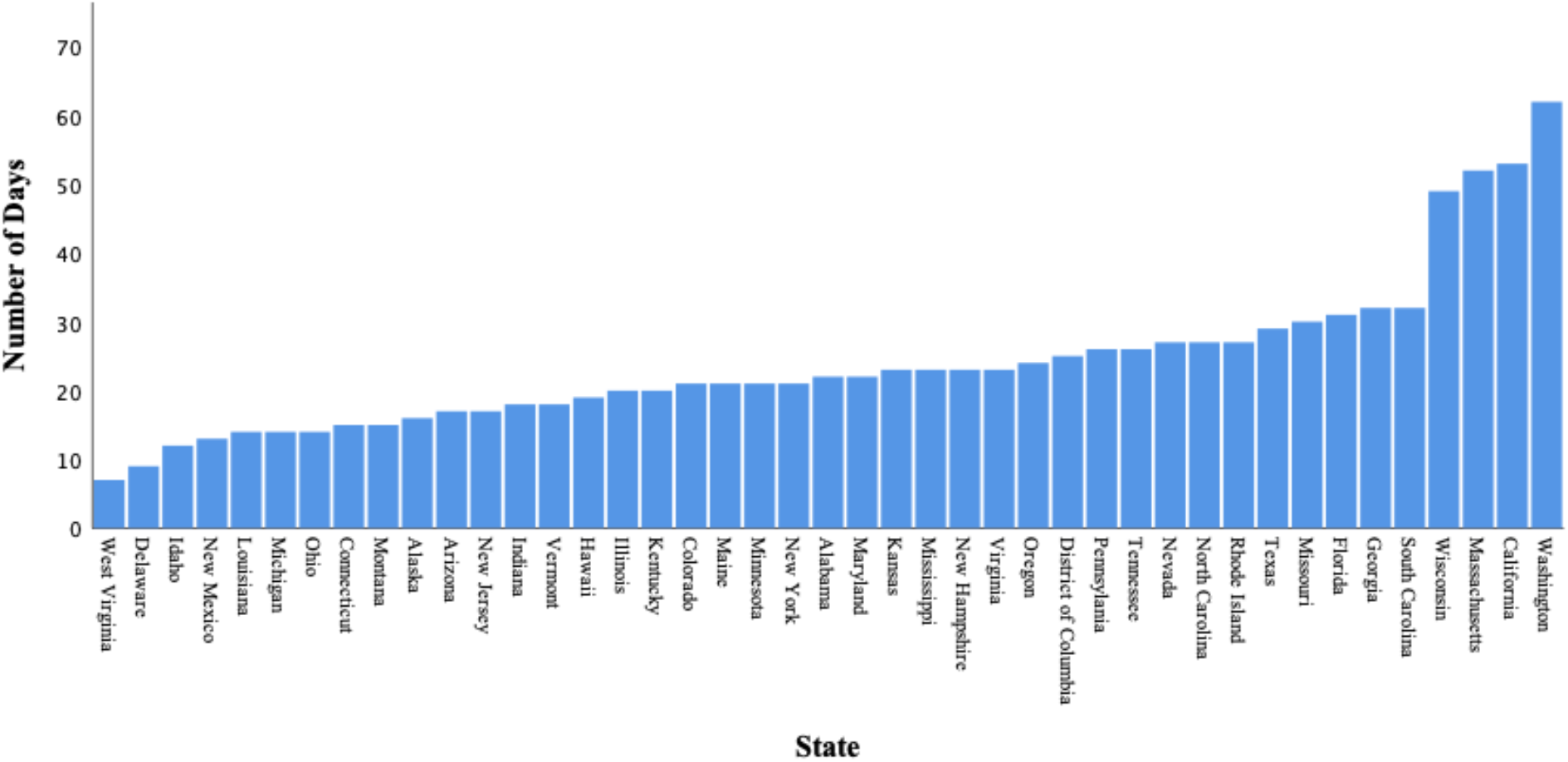
Number of Days Between Date of First Reported Case and Stay-at-Home Mandate per US state (n=43)

**Figure 2:**
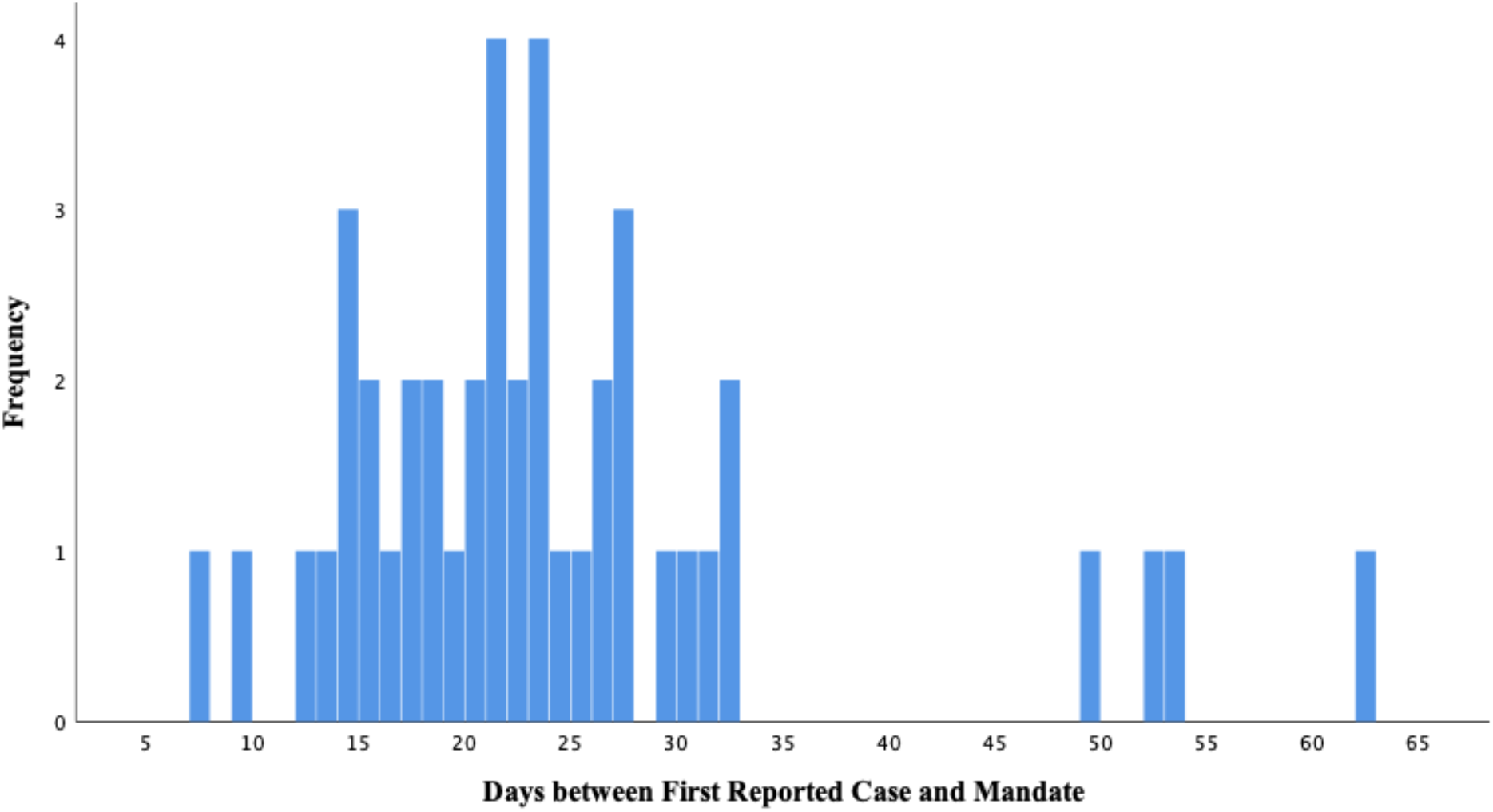
Distribution of the Number of Days Between Date of First Reported Case and Stay-at-Home Mandate per US state (n=43)

### Country-Level Descriptive Analysis

Forty-one countries with stay-at-home orders were included in our analysis. Of the 41 countries included, the number of days between the first reported case and the stay-at-home mandate ranged from 5 to 59 days (Fig. 3), with a mean of 25.2 days and a standard deviation of 14.9 days (Fig. 4).

**Figure 3:**
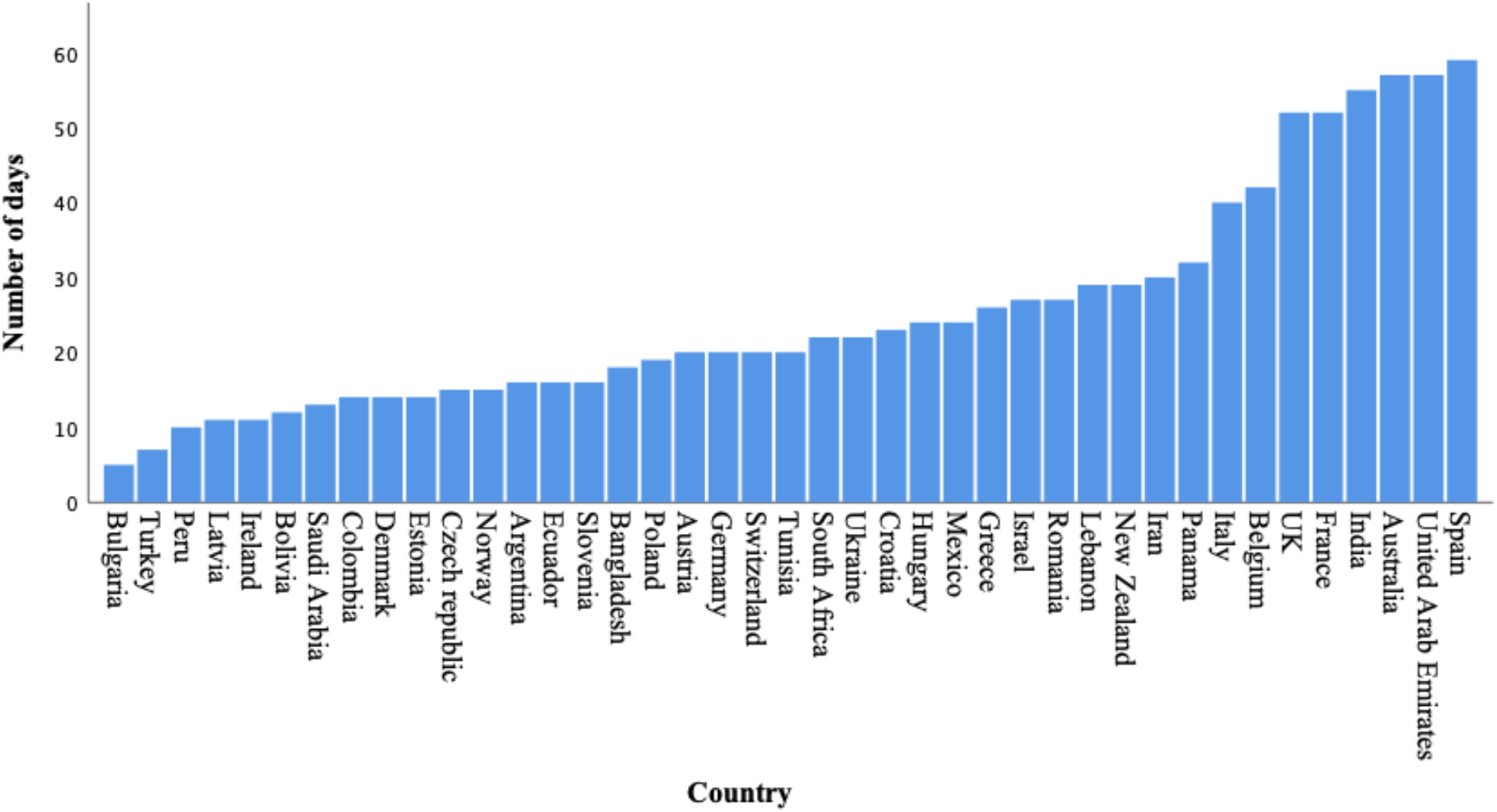
Number of Days Between Date of First Reported Case and Stay-at-Home Mandate per Country (n=41)

**Figure 4:**
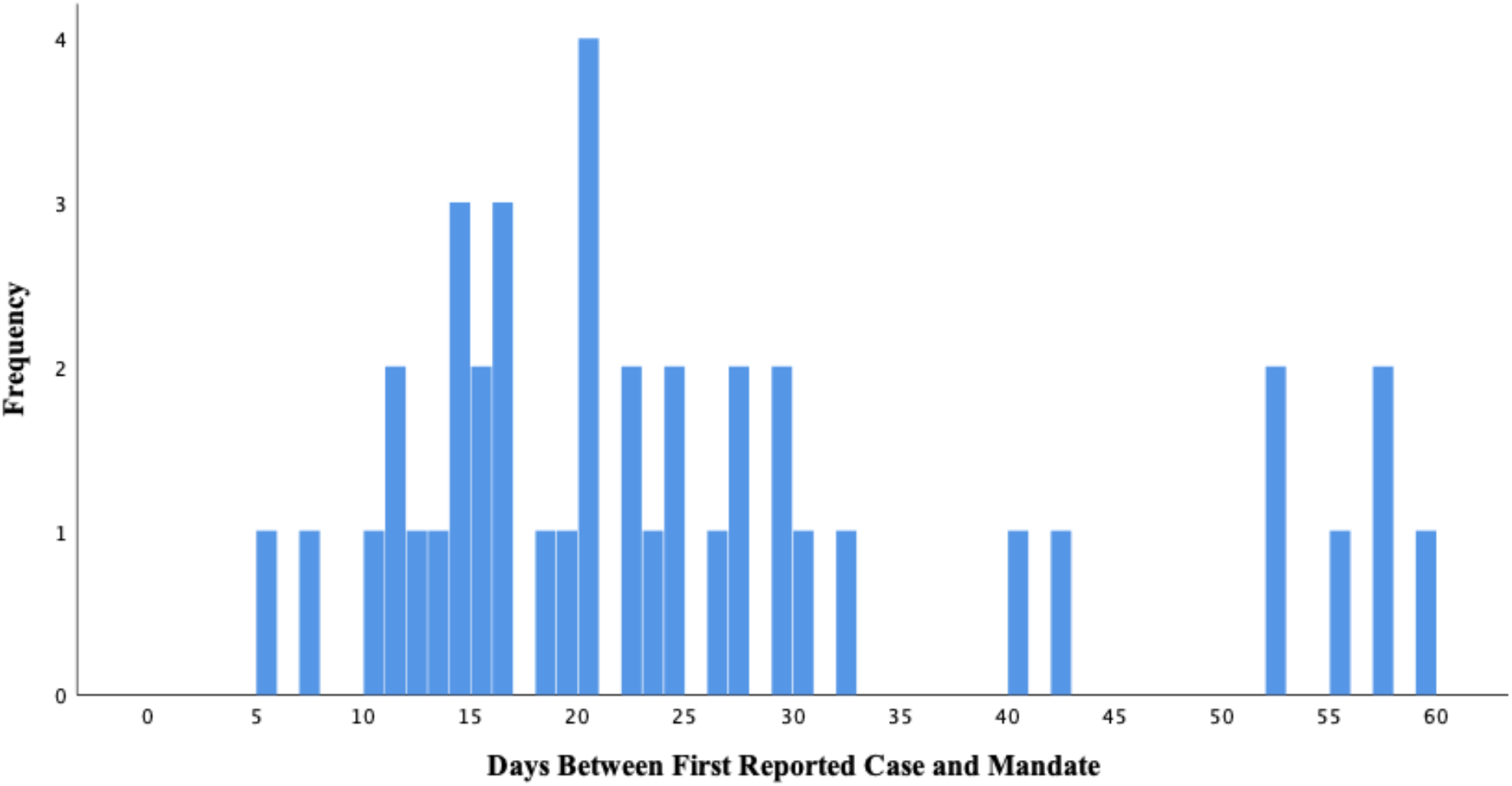
Distribution of the Number of Days Between Date of First Reported Case and Stay-at-Home Mandate per Country (n=41)

### Linear Regression Analysis

For both our country and US state-level-data, a larger number of days between the first reported case and the stay-at-home mandate was associated with a longer time to reach both the peak of daily cases and deaths for each respective region (Tables 1a and 1b). For the US states, each additional day added between the first reported case and the implementation of a mandate predicted an extra 1.1 days to reach the peak number of cases (95 % CI: 0.7, 1.5) and an extra 1.0 days to reach the peak number of deaths (95 % CI: 0.7, 1.4). The largest effect was among regions classified as the latest 10% to implement a mandate, which in the US, predicted an extra 35.3 days to the peak number of cases (95 % CI: 18.2, 52.5), and 38.3 days to the peak number of deaths (95 % CI: 23.6, 53.0). A protective effect was not seen for the countries and states that were identified as the earliest 10% of regions to implement their mandates, respectively. Classifying states and countries into categorical terciles yielded mixed results, elucidating stronger associations for state-level compared to country-level data.

**Table 1a.**
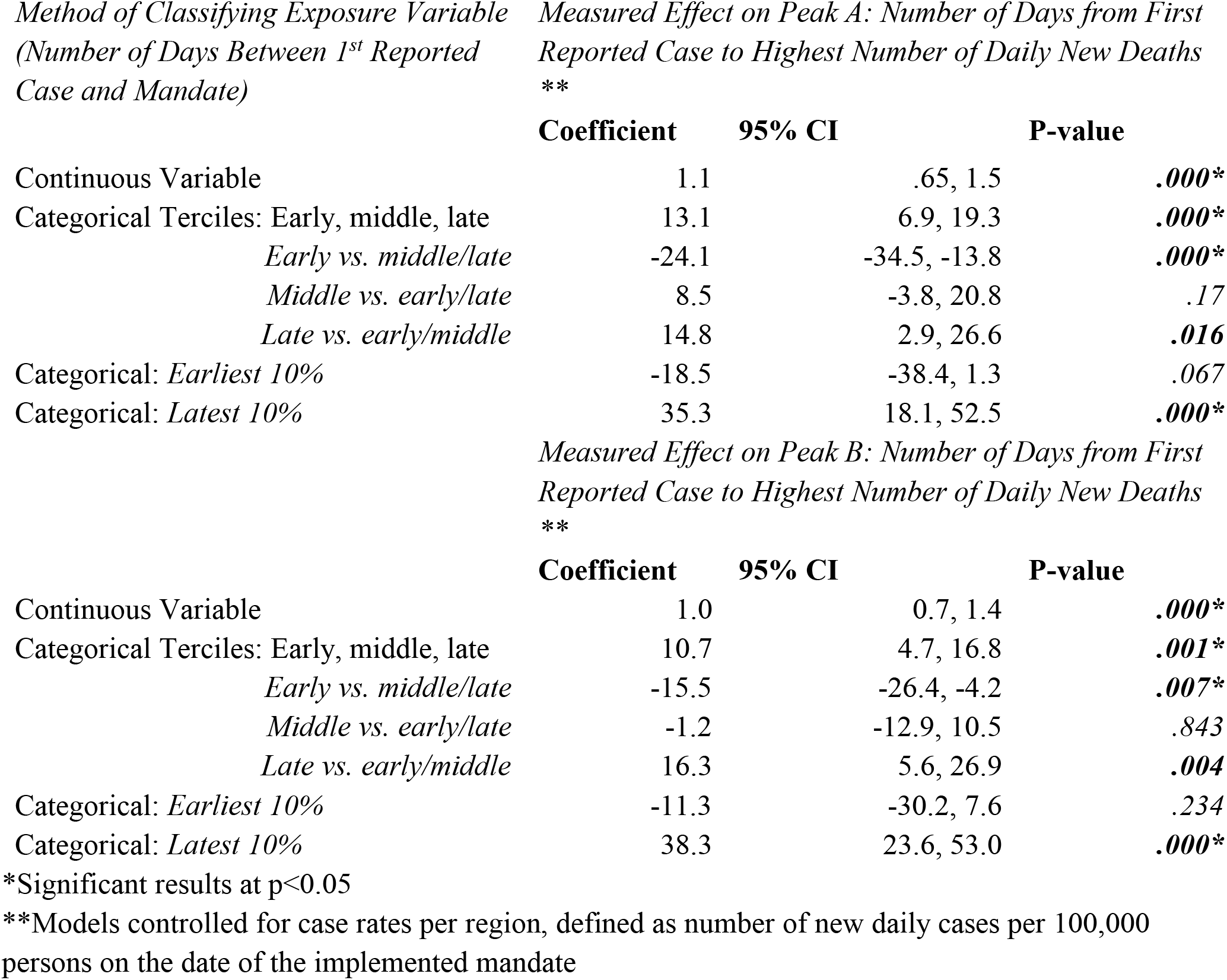
Linear Regression Models Predicting Number of Days to Highest Case and Death Count for State-level Analysis (n=43)

**Table 1.**
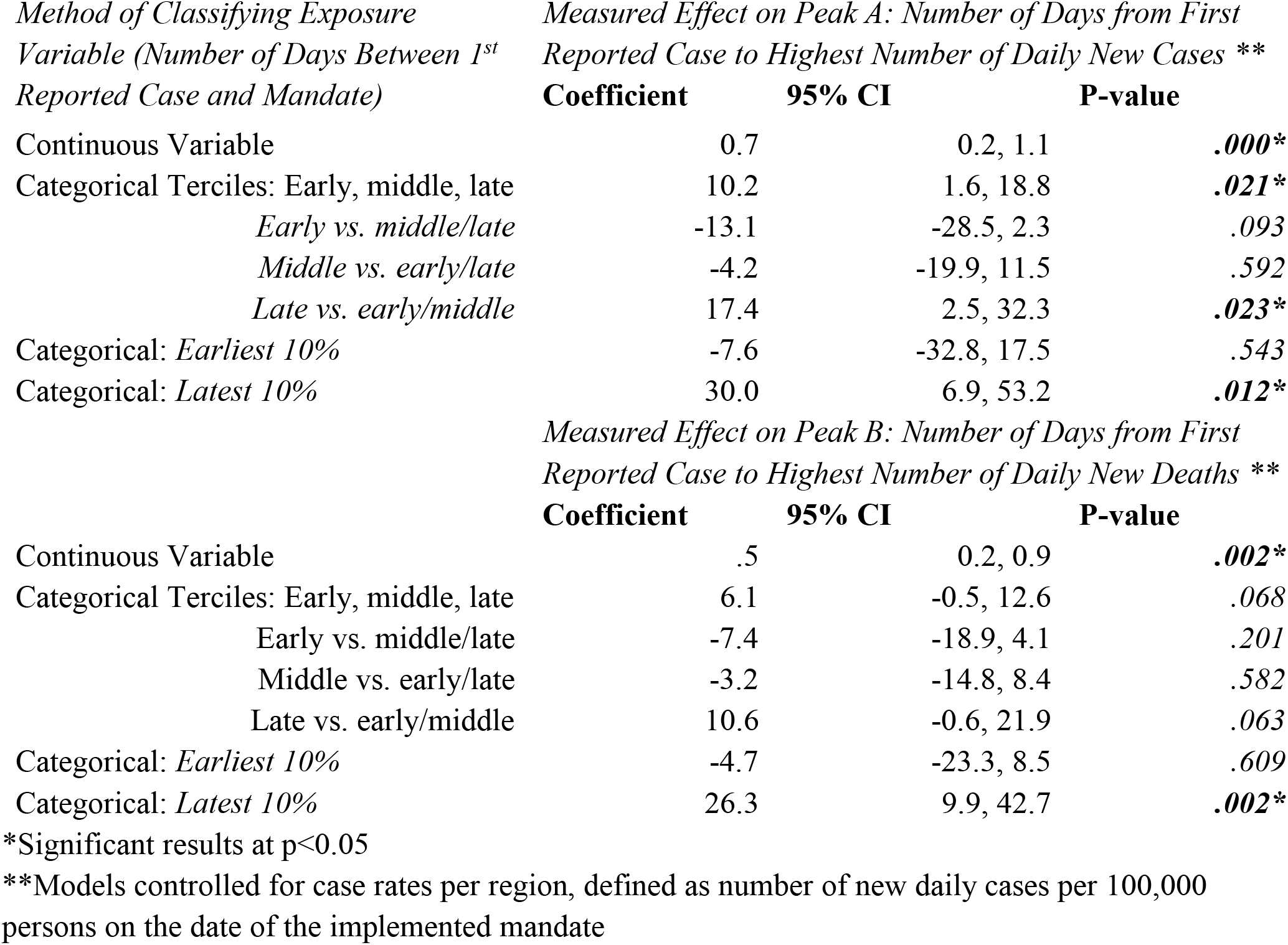
Linear Regression Models Predicting Number of Days to Highest Case and Death Count for Country-level Analysis (n=41)

## Discussion

Our study builds on the recent emerging epidemiological data supporting the efficacy of NPIs, and specific to our study, stay-at-home mandates, in the control of the COVID-19 pandemic.^6,17,16,18-22^ Our analysis supports a protective effect of earlier implementation of stay-at-home orders both globally and within US states. Notably, when the timing of mandate implementation was analyzed as a continuous variable, the effect on timing to peak case and death counts was modest with an increase in the time to peak of approximately one day. By contrast, a relatively strong effect was demonstrated when we evaluated regions categorized as late mandate implementers, corresponding to the largest predicted prolongation in the number of days to peak daily case and death counts. This strong association supports the possibility of a “threshold” date or range of dates only until which an implemented mandate may be efficacious.

Strengths of this study include the temporality of the interventions and outcomes included in our analysis, which supports biological plausibility. Furthermore, our study included multiple iterations of analyses to support the observed trend. Our findings were replicated for both US states as well as for our included countries, which supports the consistency of the observed effect. Finally, we accounted for the relative burden of disease at the time of each region’s mandate, by controlling for the case rate of disease for each country and US state included in our regression models.

The main limitation of this study was its observational nature and the exclusion of other NPIs, possibly confounding, that were implemented in the various regions we analyzed. However, we assume that by virtue of including many different regions and by repeating our analysis in several different ways, we can assume that the overall preventative effect of these NPIs were evenly spread out across these regions.^22^ Furthermore, another limitation of our study is that we did not account for the fidelity of and adherence to the implemented mandates which may have therefore biased our results. However, the directionality of this bias is unknown. Finally, the differences between regions as well as changes in testing capacity within each respective region may have also largely impacted the results of this study, as alluded to in other epidemiological observational studies that have recently investigated this topic.^6,18^

Overall, our study supports the potential effect of earlier stay-at-home mandates in the control of the spread of COVID-19. While this effect was modest generally, regions that significantly delayed implementation of their stay-at-home mandates experienced a pronounced and prolonged delay in reaching both peak daily case and death counts of COVID-19.

## Data Availability

Data received from public health websites, WHO situation reports, New York Times, worldometer.com

